# Targeting Versus Tailoring Educational Videos for Encouraging Deceased Organ Donor Registration in Black-Owned Barbershops

**DOI:** 10.1101/2021.10.28.21265263

**Authors:** Stephen P. Wall, Patricio Castillo, Francine Shuchat-Shaw, Elizabeth Norman, David Brown, Natalia Martinez-López, Mairyn López-Ríos, Azizi A. Seixas, Jan L. Plass, Joseph E. Ravenell

## Abstract

In the U.S., Black men have highest risk for requiring kidney transplants but are among those least likely to register for organ donation. Prior outreach used videos culturally targeted for Black communities, yet registration rates remain insufficient to meet demand. Therefore, we assessed whether generic versus videos culturally targeted or personally tailored based on prior organ donation beliefs differentially increase organ donor registration. In a randomized controlled trial, 1,353 participants in Black-owned barbershops viewed generic, targeted, or tailored videos about organ donation. Logistic regression models assessed relative impact of videos on: 1) immediate organ donor registration, 2) taking brochures, and 3) change in organ donation willingness stage of change from baseline. Randomization yielded approximately equal groups related to demographics and baseline willingness and beliefs. Neither targeted nor tailored videos differentially affected registration compared with the generic video, but participants in targeted and tailored groups were more likely to take brochures. Targeted (OR=1.74) and tailored (OR=1.57) videos were associated with incremental increases in organ donation willingness stage of change compared to the generic video. Distributing culturally targeted and individually tailored videos increased organ donor willingness stage of change among Black men in black-owned barbershops but was insufficient for encouraging registration.

---

Over 90,000 patients are waitlisted for kidney transplants in the U.S. (OPTN Data, 2021). Black Americans comprise 32% of the waitlist and 13% of the overall population compared with non-Latinx Whites comprising 35% and 76% respectively (U.S. Census Bureau, 2019). Because Black Americans have higher prevalence of hypertension and diabetes, two leading causes of kidney failure, this disparity persists without signs of mitigating (U.S. HSS, 2020). The disparity is even more pronounced among Black men who comprise 19% compared with Black women who comprise 13%.

Enrolling more Black Americans in organ donor registries may alleviate the disparity. After states established registries, donations increased by 8% with 33% enrollment among eligible donors (Callison & Levin, 2016). Despite 85% of Black Americans supporting organ donation, only 38% register − the lowest rate among all racial/ethnic groups, compared with ∼60% national average (Donate Life America, 2019; HRSA, 2019). In NY State, registration rates are even lower. Only 28% of New Yorkers register contributing to 15% enrollment among eligible donors (Donate Life America, 2019; U.S. Census Bureau, 2019).

Prior outreach encouraged registration in schools, colleges, universities, businesses, clinical, and community settings − often via video, multimedia, and social media (K. R. J. Arriola et al., 2019; Callender & Miles, 2010; Deedat, Kenten, & Morgan, 2013; D. DuBay et al., 2020; Quick, King, Reynolds-Tylus, & Moore, 2019; Resnicow et al., 2010; Rodrigue, Boger, DuBay, & Fleishman, 2019; Rodrigue, Fleishman, Fitzpatrick, & Boger, 2015; Steenaart, Crutzen, Candel, & de Vries, 2019; Thornton et al., 2019; Thornton et al., 2016). The overwhelming majority of registrations (93%) occurs at Departments of Motor Vehicles (DMVs) (HRSA, 2019). Reasons include: i) designation is legally represented on driver licenses, so motorists expect to discuss their wishes during DMV transactions, and ii) DMVs offer immediate opportunities to register (Alvaro, 2011; HRSA, 2019; J. T. Siegel et al., 2010). Consequently, many outreach programs prioritize DMVs (D. DuBay et al., 2020; Quick et al., 2019; Rodrigue et al., 2015; Thornton et al., 2012).

Unfortunately, in dense cities many persons lack driver licenses (e.g., only 46% licensed in NYC) (NY DMV, 2018; U.S. Census Bureau, 2019). Black-owned barbershops (BOBs) are potentially favorable alternative settings for promoting organ donation (Linnan, D’Angelo, & Harrington, 2014). BOBs are popular sites where Black men of all socioeconomic strata gather frequently and feel comfortable discussing details of their lives, including important health issues (Murphy, 1998). Also, Black barbers are influential peers with a long history of shaping public opinion (Ferdinand, 1995, 1997; Mitka, 2004; Murphy, 1998). On average, clients get haircuts every week or two from the same barber in a relationship that often lasts >10 years (Hess et al., 2007). BOBs have been beneficial for promoting cardiovascular health and cancer prevention (Linnan et al., 2014; Victor et al., 2018; Victor et al., 2011). Organ donation outreach is a natural extension of health promotion in BOBs because Black men are among those most likely to require transplants (Donate Life America, 2019; HRSA, 2019).

BOBs are suitable for showing videos that not only are culturally targeted but personally tailored to baseline organ donation beliefs. Most standard educational programming about organ donation presents generic content appealing to broad audiences, yet prior research suggests messaging targeted to beliefs shared among Black communities improves willingness to donate organs over generic content (Andrews et al., 2012; K. R. J. Arriola et al., 2019; Callender & Miles, 2010; Deedat et al., 2013; D. A. DuBay et al., 2017; D. A. DuBay et al., 2018; Morgan & Cannon, 2003; Siminoff & Sturm, 2000). Personally tailoring interventions has shown further improvements over cultural targeting to encourage cancer screening, nutrition, tobacco cessation, and kidney donation from living donors (Bull, Kreuter, & Scharff, 1999; Kreuter, Bull, Clark, & Oswald, 1999; Kreuter, Caburnay, Chen, & Donlin, 2004; Kreuter, Lukwago, Bucholtz, Clark, & Sanders-Thompson, 2003; Kreuter, Skinner, et al., 2004; Kreuter & Strecher, 1996; Nansel, Weaver, Jacobsen, Glasheen, & Kreuter, 2008; Waterman et al., 2014; Wirken et al., 2018). The key difference between cultural targeting and personally tailoring is tailoring requires assessment of an individual’s baseline beliefs (e.g., using a survey), and then the intervention is personalized for optimal impact. With targeting, a culturally appropriate “one-size-fits-all” intervention is allocated (Kreuter et al., 2003).

Our research goal was to assess whether generic versus culturally targeted or personally tailored educational videos, produced for Black men in BOBs, differentially affect organ donor registration immediately after viewing. Secondary outcomes included taking informational brochures and changes in organ donation willingness stage of change (ODWS) from baseline. We hypothesized that culturally targeted and personally tailored videos would increase registration, taking brochures, and ODWS compared with generic videos, with personally tailored videos performing best.

## Method

### Study design/setting

The randomized controlled trial occurred in 41 BOBs throughout Harlem and Brooklyn, NYC, from May 2015-November 2017. The project manager and one research coordinator were Black men who collaborated previously with BOBs throughout NYC to encourage hypertension and colorectal cancer screening (Cole et al., 2017). The other two coordinators were women from Puerto Rico. Team members lived within and were deeply familiar with the communities where BOBs were located.

Research coordinators surveyed neighborhoods to locate BOBs and spoke with owners about participation. BOBs were recruited if the owner(s) self-identified as Black and client volumes and space were anecdotally sufficient for participation. Owners received $200 if recruiting quotas (25 participants) were met.

### Participants

Participants were Black men ≥18 years old who spent ≥28 consecutive days annually in NY, New Jersey (NJ), or Connecticut (CT), spoke English, and understood consent processes. Exclusion criteria were prior registration, visual/hearing impaired, or refused.

### Video interventions

Educational-entertainment models (Supplement-A), grounded in cognitive science, education, and social psychology informed the design of videos that would motivate viewers to live vicariously through testimonials and recall prior experiences, all to encourage registration (Singhal, Cody, Rogers, & Sabido, 2004). Social cognitive learning theory suggests “modeling” realistic situations, social relationships, and attitudes with character portrayals of both careless and healthy behaviors and their consequences (Bandura, 1986). Cognitive dissonance suggests media designs that juxtapose multiple opposing perspectives induce cognitive conflict, thereby heightening engagement and problem-solving (Elliot & Devine, 1994). Anchored instruction proposes portraying meaningful problematic social situations presented with multiple perspectives and alternative solutions through character interactions with unresolved endings that challenge viewers to explore problems further (CTGV, 1990).

To further inform content, we conducted ∼30 min semi-structured interviews with 77 Black men in four BOBs purposefully recruited to have representation of those unwilling, undecided, and willing to register (Tables-S1&S2). Qualitative analysis revealed religious, cultural/knowledge, altruistic, and normative (perceptions of family and friends) beliefs influenced registration decisions (Radecki & Jaccard, 1997) (Supplement-B). Salient themes were: 1) Black men who expressed willingness to donate organs believed the physical body is not required for the afterlife, and the organ donation system prioritizes those with greatest need; 2) those unwilling mistrusted the medical system, believed transplants are reserved for wealthy patients, White patients are prioritized, and wealth supersedes race for allocation decisions; 3) participants would consider registering if they could select deserving recipients; 4) participants preferred real testimonials over acted portrayals (Supplement-B & Table-S3). These findings corroborated other research conducted within Black communities (D. A. DuBay et al., 2017; Morgan & Cannon, 2003; Siminoff & Sturm, 2000). Guided by our educational-entertainment experts (Supplement-A), a professional health media company (Imagine Health http://imaginehealth.squarespace.com/) produced the 4-6 min educational videos.

### Generic video

*Kelly’s Story* portrays standard uplifting content for diverse audiences with White protagonists (Wall, Nijhon, & Kauffman, 2012). It begins with Mary describing her daughter Kelly’s devastating head injury sustained while hiking and how organ donation specialists requested authorization compassionately. Michael, the recipient of Kelly’s heart, and his wife describe their struggles from waiting. Black, Latinx, and White experts from NYC describe how organ shortages cause thousands of unnecessary deaths annually and the disproportionate need in vulnerable communities. Experts debunk myths, focusing on domains from the Behavioral Model for Organ Donation (Radecki & Jaccard, 1997). Afterwards, Mary and Michael embrace tearfully when meeting for the first time. The video concludes when Mary encourages viewers to register stating, “You can do this.”

### Targeted video

*James’ Story* (Wall, Nijhon, & Castillo, 2015), was scripted according to education-entertainment models, so clients would live vicariously through the plight of kindred spirits (Kincaid, 2002; Singhal et al., 2004). The video begins at Denny Moe’s Superstar Barbershop in Harlem. Denny and two clients, an older Black woman and a young Black man, discuss beliefs shared from the formative interviews. They acknowledge organ donation “is not something they really think about,” and that “We don’t trust going to the hospital and letting people take things out of us.” Differing perspectives are shared by clients in BOBs to induce cognitive conflict among viewers, thereby heightening engagement and problem-solving (Elliot & Devine, 1994). James discusses his struggle to support his family while waiting for >4 years for kidney transplantation. Intermixed are uplifting scenes with his daughters and emotional scenes with his partner lamenting, “The longer he lives without a kidney, he’ll be declining slowly…it’s hard,” with James responding, “You know, I never stopped to think about it from your side.” Respected barbers recall stories of friends who died while waiting. Black doctors and a Reverend debunk misconceptions shared from clients, organized by religion, culture/knowledge, altruism, and normative beliefs. Returning to Denny’s barbershop, the young Black man states he would consider signing up. The video ends when James ruminates, “Hopefully someone, somewhere will help me…I just patiently wait and wait.” The unresolved ending leaves viewers to imagine the outcome that they hope for, expect, or fear for James and the consequences (Singhal et al., 2004).

### Tailored videos

Prior research suggests learners perform better if content is presented at their level of understanding, rather than having to filter out known content from the unknown (Homer & Plass, 2010; Kalyuga, 2009). Therefore, producers systematically cut religious, culture/knowledge, altruistic, and normative content from *James’ Story* until all 16 possible combinations were produced (Table-1). Videos shared introduction, following of James’ struggles, and ending scenes. Using a validated organ donation belief index (ODBI Table-S4) (Wall et al., 2010), we generated belief profile signatures at baseline for each participant using the following algorithm: if a participant scored ≥.6 logits in a domain, that content was deemed redundant and removed from presentation. For example, a participant who scored 1.1 religion, .2 culture/knowledge, 1.6 altruism, and .4 normative, profiled as “CN” and viewed the video having culture and normative content included and religious and altruism content excluded. Participants who scored <0.6 logits for all domains profiled as “RCAN” and viewed the complete video.

**Table 1:** Allocation of Videos among Participants by Idiographic Profile.

|  | Tailored<br>N = 456 | Targeted<br>N = 450 | Generic<br>N = 447 |
| --- | --- | --- | --- |
| **Film Viewed** | **n (%)** |  |  |
| Generic Film |  |  | 447 (100) |
| Complete Film | 218 (47.8) | 450 (100) |  |
| RCN | 104 (22.8) |  |  |
| RC | 25 (5.5) |  |  |
| CN | 21 (4.6) |  |  |
| RN | 20 (4.4) |  |  |
| C | 17 (3.7) |  |  |
| RAN | 16 (3.5) |  |  |
| RCA | 16 (3.5) |  |  |
| N | 6 (1.3) |  |  |
| CAN | 4 (0.9) |  |  |
| R | 3 (0.7) |  |  |
| Only the Story | 2 (0.4) |  |  |
| A | 1 (0.2) |  |  |
| AN | 1 (0.2) |  |  |
| CA | 1 (0.2) |  |  |
| RA | 1 (0.2) |  |  |

R – religion, C- culture/knowledge, A – altruism; N – normative. Content for each domain was shown if the standardized score for each domain was < 0.6 logits. If participants scored <0.6 logits in all domains, the complete film was shown, the same film that was allocated to participants in the targeted group.

### Measures and outcomes

The primary outcome was immediate enrollment in NY, NJ, or CT Organ Donor Registries. Secondary outcomes included taking informational brochures, ODWS (Table-S5) (Prochaska, Redding, & Evers, 2015); ODBI scores for religious, culture/knowledge, altruism, and normative domains; and an intrinsic motivation inventory (IMI Table-S6). ODBI scores were measured with 20 items and IMI with 12 items using 7-pt bipolar responses administered with branching logic (Malhotra, Krosnick, & Thomas, 2009; McAuley, Duncan, & Tammen, 1989). Participant demographics included age, education, religion, and ethnicity.

### Protocol

Candidates were screened in BOBs. For those eligible and who agreed, research coordinators conducted baseline assessments, presented videos on iPads with noise cancelling headphones, and conducted post-assessments. If participants wanted to register, research coordinators assisted with completing enrollment forms, after which forms were mailed in preaddressed, stamped envelopes. Assessments, random allocation in blocks of 12, and video distribution was facilitated with SherlockMD© Clinical Research Software (Santa Monica, CA). Before participants viewed videos, research coordinators stepped away to maintain blinding to assignment. Participants provided verbal informed consent for participation without incentives to avoid coercion. Our institutional review board approved the research. A data and safety monitoring board ensured participant safety, and a community advisory board, comprised of barbershop owners, activists, and clergy, advised on recruitment and video production (Supplement-C).

### Analysis

Categorical outcomes were assessed with proportions and confidence intervals (CIs) stratified by video assignment and adjusted for clustering within BOBs. We tested inequalities among treatment groups using multilevel logistic regression. Registration was the dependent variable, video group was the primary regressor, and BOB location was the random effect (Campbell, 2000). We used ordinal logistic regression to model ODWS. We calibrated the ODBI and IMI with multi-dimensional random coefficient multinomial logit modeling using ACER ConQuest 4.0 (R. Adams, Wu, & Wilson, 2015; R. J. Adams, Wilson, & Wang, 1997). Expected a-posterior ability estimates were calculated for each belief domain. Linear regression with post-intervention ability as the dependent variable, video group as primary regressor, and pre-intervention ability as a covariate was modeled separately for each domain adjusting for BOB clustering. Outcomes were assessed primarily with intention to treat and secondarily as treatment effects.

When >5% of data were missing for an estimated variable, data analyses were performed by combining model estimates run on 20 multiply imputed data sets using Rubin’s rules (Rubin & Schenker, 1991). For the ODBI and IMI calibrations, refused to answer, not applicable, or missing responses were coded as missing, with imputations undertaken using plausible values (Mislevy, 1991). We assessed logistic regression model fit with Hosmer-Lemeshow tests and proportional odds assumptions for ordinal logistic regressions with Brant tests (Brant, 1990; Hosmer Jr, Lemeshow, & Sturdivant, 2013). We examined normality assumptions in the linear regression models with kernel density plots of predicted versus actual residuals and Breusch and Pagan Lagrangian multiplier tests for random effects (Breusch & Pagan, 1980). Analyses were performed with Stata version 16 (College Station, TX).

Prior research conducted in BOBs revealed an intra-cluster correlation coefficient of .044 (Victor et al., 2011). To assure power above .90, with significance level of .017 (correction for three comparisons), required at least 1,350 participants from 27 BOBs (450 in each arm) with an effective sample size of 50 participants per cluster. Power calculations were based on preliminary data suggesting baseline willingness to register rate of 15% among Black men in the generic arm and improvement to 35% in the experimental arms, using a .65 coefficient of variation. We used fitted-models to compute intra-cluster correlations. Adjustments were made if recruiting targets within each BOB were unmet, by increasing the number of clusters and reducing recruitment per cluster to achieve similar power. Power calculations were accomplished using PASS (Kaysville, Utah).

## Results

### Study population

Research coordinators approached 3,748 Black men in 41 BOBs (Figure-1). Main reasons for exclusion were prior registration (14%) and refusals (73%). When approached, 1,440 (38%) participated with 1,353 (94%) completion. Dropouts included equipment failures and left before completion. Mean age was 34 years; 15% self-identified as Black-Latinx, 69% had education beyond high school; 60% were Christian, 6% Islamic, and 34% agnostic, atheist, or practicing other religions. Randomization yielded groups balanced on age, ethnicity, education, religion, baseline ODWS and baseline ODBI scores (Table-2).

**Table 2.** Baseline Demographic Characteristics, Willingness, and Beliefs.

|  | **Generic**<br>**N = 447** | **Targeted**<br>**N = 450** | **Tailored**<br>**N = 456** |
| --- | --- | --- | --- |
|  | **u (Std)** | **u (Std)** | **u (Std)** |
| **Age** | 33.5 (11.8) | 34.1 (12.0) | 33.6 (11.6) |
| **Ethnicity** | **n (%)** | **n (%)** | **n (%)** |
| Non-Latino | 388 (86.8) | 380 (84.4) | 386 (84.6) |
| Latino | 59 (13.2) | 70 (15.6) | 70 (15.4) |
| **Place of Birth** |  |  |  |
| US | 370 (82.6) | 370 (82.0) | 375 (82.0) |
| Outside US | 77 (17.4) | 80 (18.0) | 81 (18.0) |
| **Education** |  |  |  |
| < High School | 23 ( 5.2) | 16 ( 3.6) | 19 ( 4.2) |
| High School or GED | 112 (25.1) | 126 (28.0) | 126 (27.6) |
| Some College | 118 (26.4) | 118 (26.2) | 104 (22.8) |
| Associate | 48 (10.7) | 48 (10.7) | 38 ( 8.3) |
| Bachelors | 100 (22.4) | 94 (20.9) | 121 (26.6) |
| Graduate Education | 46 (10.3) | 48 (10.7) | 48 (10.5) |
| **Religion** |  |  |  |
| Catholic | 50 (11.2) | 45 (10.0) | 47 (10.3) |
| Christian Not Specified | 226 (50.6) | 211 (46.9) | 227 (49.8) |
| Islam | 25 ( 5.6) | 31 ( 6.9) | 21 ( 4.6) |
| Agnostic or Atheist | 18 ( 4.0) | 18 ( 4.0) | 23 ( 5.0) |
| Other<sup>2</sup> | 128 (28.6) | 145 (32.2) | 138 (30.3) |
| **Organ Donor Willingness** | **n (%)** | **n (%)** | **n (%)** |
| I Do Not Want to Sign Up | 134 (30.0) | 120 (26.7) | 145 (31.8) |
| I Do Not Think I Want to Sign Up | 82 (18.3) | 88 (20.0) | 87 (19.1) |
| I May Want to Sign Up | 186 (41.6) | 199 (44.2) | 185(40.6) |
| I Would Like to Sign Up | 37 (8.2) | 27 (6.0) | 23 (5.0) |
| I Definitely Want to Sign Up | 8 (1.8) | 16 (3.6) | 16 (3.5) |
| **Organ Donation Belief Scores<sup>2</sup>** | **u (Std)** | **u (Std)** | **u (Std)** |
| Religion | 0.04 (0.037) | 0.06 (0.036) | -0.02 (0.031) |
| Culture/Knowledge | -0.21 (0.030) | -0.12 (0.031) | -0.23 (0.031) |
| Altruism | 0.47 (0.050) | 0.60 (0.050) | 0.49 (0.041) |
| Normative | -0.08 (0.050) | 0.02 (0.053) | -0.11 (0.036) |

All participants self-identified as Black men.
N – number of participants within a treatment group; n – number of participants within each cell; u – mean; Std – standard deviation
<sup>1</sup>Other categories include other religions not classified as Christian (i.e. Judaism), and refused to answer.
<sup>2</sup>Scores for Religion, Culture/Knowledge, Altruism, and Normative constructs from the Organ Donation Belief Model were calibrated with Rasch analyses using the multidimensional partial credit model in ACER ConQuest Software version 4.0 (Sydney, Australia). Instrument items are presented in Tables S4 & S6. Means reported using multiple imputation on 20 repeated data sets sampled with replacement and combined using Rubin's rules. Std was adjusted for clustering at the Barbershop level.

**Figure 1:**
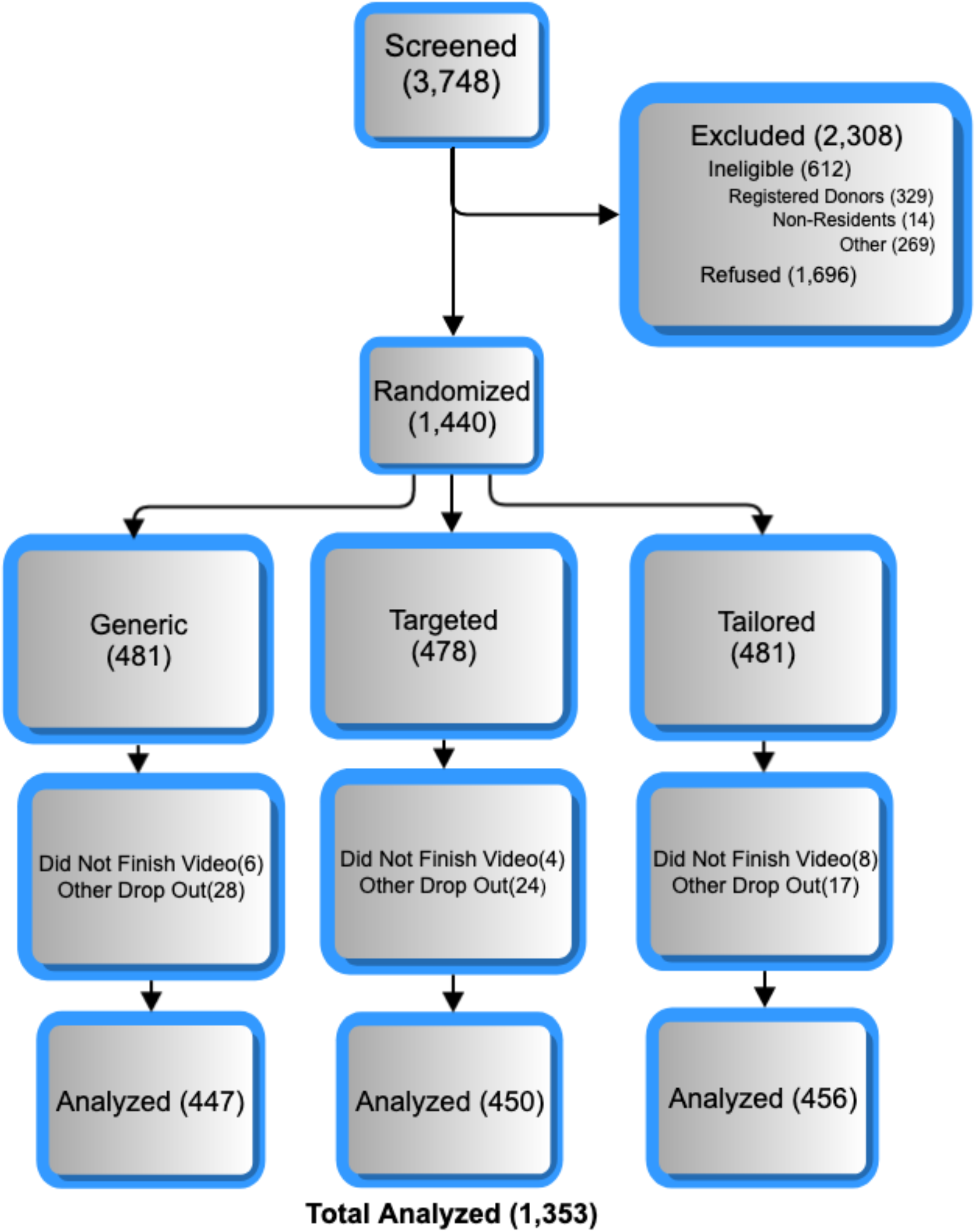
CONSORT Diagram.

### ODBI and IMI Calibrations

ODBI deviances for rating scale (135924) and partial credit (134189) models were significantly greater than multi-dimensional model deviance (129520; *p* < .001). Questions mapped to four dimensions (Table-S4). Domains included religion (5-questions), culture/knowledge (7-questions), altruism (4-questions), and normative (4-questions). Overall separation reliability was 1.00. Questions exhibited weighted mean squares .83–1.40. Question-5, despite having weighted mean square >1.33 indicating less than optimal discrimination (Smith Jr, 2000), was retained because it was used in the personalization algorithm. Intrinsic motivation inventory (IMI) scale deviances for rating scale (16486) and partial credit (16477) models were significantly greater than multi-dimensional model deviance (15488; *p* < .001). Responses were merged into 4 unipolar categories (Not-at-all/neutral, Believe-a-little, Believe-moderately, Believe-a-lot) because unfavorable ratings were endorsed infrequently (Table-S6). Questions mapped to two dimensions after removing 2 underperforming questions. Domains included enjoyment (6 questions) and value (6 questions). Overall separation reliability was .986. Questions exhibited weighted mean squares .87–1.27. Both scales exhibited item total correlations >.4 and item point-biserial correlations increased for each of the response categories. Calibrations yielded standardized scores for each domain with scores falling within -4 to 4 logits and centered around 0 (Smith Jr, 2000). Higher values indicated more favorable beliefs about organ donation/intrinsic motivation.

### Intention-to-treat analyses

Overall, 10% of participants registered, and 54% took brochures. Unadjusted analyses (Table-3) revealed registration rates for the targeted (12.2%, 95%CI [9.2,16.1]) and tailored (9.2%, 95%CI [6.9,12.2]) groups were not different significantly from the generic video (9.6%; 95%CI [7.0,13.0]). Targeted (58.7%, 95%CI [52.5,64.6]) and tailored (55.9%, 95%CI [49.6,62.1]) groups trended towards taking brochures more often than the generic group (48.2%, 95%CI [42.7,53.7]), but results were not significant statistically. Post-viewing educational videos, targeted and tailored groups exhibited greater ODWS than the generic group (Figure-S1). Mean enjoyment and value scores for the targeted group were ∼.3 logits greater than the tailored group. Tailored and targeted group means for enjoyment and value were >∼.7 to ∼1 logit above the generic group. Targeted and tailored groups had mean religious scores from ∼.1 to ∼.2 logits greater than the generic group, but culture/knowledge mean scores were lower by .04 and .14 logits respectively. Only altruism mean scores exceeded .6 logits, the threshold for registering (Wall et al., 2010). Differences in pre-post intervention ODBI scores corroborate these findings (Table-S7).

**Table 3.** Unadjusted Outcomes ITT Analysis.

|  | **Generic**<br>N = 447 | **Targeted**<br>N = 450 | **Tailored**<br>N = 456 |
| --- | --- | --- | --- |
|  | **n (%) [95% CI]** | **n (%) [95% CI]** | **n (%) [95% CI]** |
| **Registered<sup>1</sup>** | 43 ( 9.6) [ 7.0, 13.0] | 55 (12.2) [ 9.2, 16.1] | 42 ( 9.2) [ 6.9, 12.2] |
| **Took Brochure<sup>1</sup>** | 208 (48.2) [42.7, 53.7] | 254 (58.7) [52.5, 64.6] | 250 (55.9) [49.6, 62.1] |
| **Organ Donor Willingness** | **n (%)** | **n (%)** | **n (%)** |
| I Do Not Want to Sign Up | 108 (24.1) | 71 (15.8) | 79 (17.3) |
| I Do Not Think I Want to Sign Up | 67 (15.0) | 51 (11.3) | 68 (14.9) |
| I May Want to Sign Up | 207 (46.3) | 242 (53.8) | 238 (52.2) |
| I Would Like to Sign Up | 46 (10.3) | 51 (11.3) | 41 (9.0) |
| I Definitely Want to Sign Up | 19 (4.3) | 35 (7.8) | 30 (6.6) |
| **Organ Donor Belief Scores<sup>2</sup>** | **u (Std)** | **u (Std)** | **u (Std)** |
| Religion | 0.19 (0.047) | 0.38 (0.053) | 0.26 (0.039) |
| Culture/Knowledge | 0.11 (0.041) | 0.07 (0.049) | -0.05 (0.043) |
| Altruism | 0.75 (0.059) | 0.94 (0.047) | 0.76 (0.048) |
| Normative | 0.10 (0.054) | 0.26 (0.063) | 0.09 (0.040) |
| **Intrinsic Motivation Scores<sup>2</sup>** | **u (Std)** | **u (Std)** | **u (Std)** |
| Enjoyment | 1.14 (0.12) | 2.15 (0.11) | 1.85 (0.13) |
| Value of Video | 0.64 (0.13) | 1.82 (0.12) | 1.49 (0.12) |

N – number of participants within a treatment group; n – number of participants within each cell; u – mean; Std – standard deviation
<sup>1</sup>Confidence intervals for proportions adjusted for clustering at the level of Barbershops.
<sup>2</sup>Scores for Religion, Culture/Knowledge, Altruism, and Normative constructs from the Organ Donation Belief Model were calibrated with Rasch analyses using the multidimensional partial credit model in ACER ConQuest Software version 4.0 (Sydney, Australia). Means reported using multiple imputation on 20 repeated data sets sampled with replacement and combined using Rubin's rules. Std was adjusted for clustering at the level of Barbershops.

Results from multivariable logistic, ordinal logistic, and linear regressions were consistent with bivariable analyses (Table-4). Neither targeted (OR=1.32, 95%CI [.86,2.03]) nor tailored (OR=1.01, 95%CI [.64,1.59]) videos differentially affected registration compared with the generic video, but targeted (OR=1.57, 95%CI [1.19,2.08]) and tailored (OR=1.31, 95%CI [1.00,1.73]) group participants took brochures more frequently. Targeted (OR=1.74, 95%CI [1.34,2.26]) and tailored (OR=1.57, 95%CI [1.21,2.03]) videos were associated with incremental increases in ODWS. Hosmer-Lemeshow GOF tests run on models not accounting for clustering indicated good model fit (p-value 1.00 for both models). Rho in models accounting for clustering within LOBs (1.41e-07) indicated negligible clustering. Brandt testing for proportional odds assumptions yielded p-Values >.05 for all parameters in unimputed models indicating ordinal logistic modeling assumptions held true.

### Treatment effects

Of 456 participants randomized to tailoring, 47.8% scored <.6 logits for all ODBI domains and subsequently watched the same video allocated to targeted group participants. We recoded these participants to the targeted group and repeated analyses. Results showed tailored videos trended towards superior impact compared with targeted videos (Tables 4&S8, Figure-S2), but differences were not significant statistically. Results confirmed no differential impact on immediate registration, regardless of video seen. Targeted and tailored videos remained significantly superior to the generic video, with participants who viewed the tailored videos having 2 times the odds of an incremental increase in ODWS compared with the generic video. When tailoring occurred, altruism content was removed most often (Table-1).

**Table 4.** Multivariable Analyses Logit & Ordinal Logit Models.

| **Categorical Outcomes** | **Intention to Treat** | **Treatment Effect** |
| --- | --- | --- |
| **Registered<sup>1</sup>** | **OR (95%CI)** | **OR (95%CI)** |
| Targeted | 1.32 (0.86, 2.03) | 1.10 (0.73, 1.66) |
| Tailored | 1.01 (0.64, 1.59) | 1.35 (0.81, 2.27) |
| **Took a Brochure<sup>1</sup>** |  |  |
| Targeted | 1.57 (1.19, 2.08) | 1.36 (1.05, 1.76) |
| Tailored | 1.31 (1.00, 1.73) | 1.66 (1.18, 2.34) |
| **Organ Donor Willingness after Viewing<sup>2</sup>** |  |  |
| Targeted | 1.74 (1.34, 2.26) | 1.54 (1.21, 1.95) |
| Tailored | 1.57 (1.21, 2.03) | 2.05 (1.49, 2.81) |
| Baseline Organ Donor Willingness | 4.88 (4.28, 5.58) | 4.86 (4.26, 5.55) |
| **Organ Donation Beliefs after Viewing<sup>3</sup>** |  |  |
| **Religion** | **Beta (95%CI)** | **Beta (95%CI)** |
| Targeted | 0.16 (0.08, 0.25) | 0.12 (0.05, 0.20) |
| Tailored | 0.12 (0.04, 0.21) | 0.20 (0.10, 0.30) |
| Baseline Religion Score | 0.81 (0.76, 0.87) | 0.81 (0.76, 0.86) |
| **Culture/Knowledge** |  |  |
| Targeted | -0.12 (-0.21, -0.04) | -0.15 (-0.22, -0.07) |
| Tailored | -0.15 (-0.23, -0.06) | -0.10 (-0.20, 0.00) |
| Baseline Culture/Knowledge Score | 0.86 (0.80, 0.92) | 0.86 (0.80, 0.92) |
| **Altruism** |  |  |
| Targeted | 0.08 (-0.01, 0.16) | 0.05 (-0.03, 0.13) |
| Tailored | -.01 (-0.10, 0.16) | -0.02 (-0.12, 0.09) |
| Baseline Altruism Score | 0.87 (0.82, 0.91) | 0.88 (0.83, 0.92) |
| **Normative** |  |  |
| Targeted | 0.08 (-0.01, 0.17) | 0.07 (-0.01, 0.15) |
| Tailored | 0.03 (-0.06, 0.11) | 0.02 (-0.09, 0.12) |
| Baseline Normative Score | 0.84 (0.80, 0.89) | 0.85 (0.81, 0.89) |
| **Intrinsic Motivation Inventory<sup>3</sup>** |  |  |
| **Perceived Enjoyment** | **Beta (95%CI)** | **Beta (95%CI)** |
| Targeted | 0.97 (0.67, 1.27) | 0.70 (0.42, 0.97) |
| Tailored | 0.69 (0.39, 0.99) | 1.19 (0.84, 1.55) |
| **Perceived Value** |  |  |
| Targeted | 1.17 (0.86, 1.51) | 0.85 (0.55, 1.15) |
| Tailored | 0.84 (0.51, 1.16) | 1.44 (1.06, 1.82) |

<sup>1</sup>Logistic regression models: registration and taking informational pamphlets were separately modelled as dependent variables with treatment groups as primary regressors (genre, ending) and LOB location as a random effect. Hosmer-Lemeshow GOF tests run on models not accounting for clustering indicated good model fit (p-value 1.00 for both models). Rho in model accounting for clustering within LOBs (1.41e-07) indicating negligible clustering effects.
<sup>2</sup>Ordinal logistic (ologit) regression models used the same strategy except the outcome was the 5pt willingness scale (see cInstruments). Brandt testing for proportional odds assumptions yielded pValues > .05 for all parameters in unimputed models indicating ologit modeling assumptions held true.
<sup>3</sup>Linear regression models: to account for missing data when >5% were missing for an estimated variable, data analyses were performed by combining model estimates run on 20 multiply imputed data sets using Rubin's rules. Normality assumptions held true for all of the linear regression models, after assessment with kernel density plots of predicted versus actual residuals and the Breusch and Pagan Lagrangian multiplier test for random effects.

## Discussion

Overall, ∼10% of participants who were unregistered for organ donation immediately registered after viewing educational videos. Although far from the 60% national rate (HRSA, 2019), this study suggests that despite collaborating with participants having low baseline willingness, outreach in BOBs can yield appreciable new registrations. This impact was similar to studies about other educational programs designed for Black communities. A study in beauty salons throughout urban areas in Michigan showed that hair stylists trained in motivational interviewing who presented comprehensive information about nutrition, exercise, diabetes, chronic kidney disease, and organ donation encouraged ∼20% of clients to register compared with 16% who received only health information (Resnicow et al., 2010). A study in churches located within Black communities throughout Southeastern Michigan showed ∼30% of participants registered after attending group sessions and watching a 32-minute video targeted for congregants compared with ∼27% from churches that did not receive interventions. Similarly, a study in Black churches throughout Atlanta compared a video targeted for congregants compared with videos and materials designed for general Black viewership; results showed no added benefit from targeted programming (K. Arriola, Robinson, Thompson, & Perryman, 2010). Furthermore, distribution of secular videos and informational materials in group sessions with participants recruited from community settings throughout Atlanta encouraged 1.3% of participants to register (K. R. J. Arriola et al., 2019).

Educational videos distributed in DMVs showed similar results compared with our study. A multicultural video presented with iPads and noise cancelling headphones led to 12% increase in registration compared with controls who received no intervention (Thornton et al., 2012). A video targeted for Black automobile drivers shown continuously in Alabama DMVs on flat screen televisions increased registration by 2.3% compared with times when the video was not shown (D. DuBay et al., 2020), and videos shown continuously in DMVs throughout Illinois and Massachusetts showed no benefit over control sites (Quick et al., 2019; Rodrigue et al., 2015). Videos that were continuously shown were inferior to distribution with iPads, likely because DMVs required silent presentation, and few drivers viewed the content (D. DuBay et al., 2020; Quick et al., 2019; Rodrigue et al., 2015).

Our results showed neither targeted nor tailored videos significantly improved registration immediately after viewing compared with generic videos. The targeted video exhibited ∼3% improvement over the generic video, and in the treatment effects analysis, the improvement shifted to the tailored videos. However, secondary outcomes support distributing targeted/tailored educational programming over generic content. Viewers of tailored and targeted videos had ∼1.5 to ∼2 times the odds of incrementally increasing ODWS and ∼1.5 times the odds of taking brochures compared with the generic video. Improving perceptions about organ donation can increase organ donation opportunities, indirectly when persons consider registration after conversations with family and peers and directly when donation decisions are made on behalf of deceased persons (Radecki & Jaccard, 1997; Siminoff, Gordon, Hewlett, & Arnold, 2001).

The improved efficacy of tailored and targeted videos was likely not from modifying organ donation beliefs. Results showed that altruism and normative beliefs did not significantly change while religious beliefs improved by only .2 logits over the generic video. Surprisingly, viewing the targeted and tailored videos lowered culture/knowledge beliefs by .1 logits compared with the generic video. The impact, though marginal, is partly explained by the targeted and tailored videos having real clients in BOBs debating content and sharing different points of view including misconceptions that are corrected by experts, whereas the generic video only had experts dispelling myths. Although education-entertainment models propose showing different points of view via conflict resolution (Elliot & Devine, 1994), when encouraging organ donor registration, our data corroborate that counterargument appeals can legitimize misconceptions (J. T. Siegel et al., 2008). Nonetheless, the targeted and tailored videos exhibited overall improvement in ODWS, likely from enjoyment/value scores significantly exceeding those from the generic video, as predicted from having Black spokespersons recounting stories and information that were more personally meaningful (Singhal et al., 2004).

Sparse attempts at tailoring educational content to encourage organ donor registration are reported. Studies from the Netherlands showed improvements in organ donation willingness among high school students using a blended video and tailored interactive survey compared with a control (Steenaart et al., 2019), but the tailored survey alone performed no better than informational brochures (Reubsaet, Brug, Kitslaar, van Hooff, & van den Borne, 2004). Tailoring occurred with an interactive survey consisting of 226 questions that pushed out text and picture messages in response to incorrect answers rather than grouping content into domains and streamlining it based on standardized scores. Equivocal results likely occurred from survey fatigue and excluding testimonials from the tailored programming (Rodrigue et al., 2019). Also, the tailored survey solely used text and still images, so it was likely less engaging than video programming.

Similarly, our results showed no significant differences in outcomes from individually tailoring compared to targeting content. We tailored the videos to remove content that was known and believed, so that participants only viewed what was new or disputed. Instead, our participants might have preferred to view content that validated some of their prior beliefs, rather than being inundated with contrary content. Tailoring underperformance also might be explained by low baseline organ donation belief scores. Only ∼50% of participants in the tailored group watched a shortened video, and most often when tailoring occurred, only altruism content was removed. Yet, another explanation is that the tailoring algorithm was suboptimal.

### Strengths and limitations

Study strengths include the randomized trial design with concealed allocation, assessing immediate registration after viewing (J. T. Siegel et al., 2010), and calibrating the ODBI and IMI using modern psychometric testing theory (R. J. Adams et al., 1997). We acknowledge pretest assessment of organ donation beliefs might have primed participants to consider organ donation, but priming was unlikely to have occurred because in prior research, pre-intervention questionnaires had no impact on modifying organ donor willingness when assessed with a Solomon four-group design (Reubsaet, Reinaerts, Brug, van Hooff, & van den Borne, 2004). This study compared the relative impact of video designs on improving organ donation registration behaviors without a control group or a group that received a sham video because a plethora of literature demonstrated that educational programs, regardless of design, improve willingness compared to doing nothing or solely reading brochures (Jason T. Siegel & Alvaro, 2010). Delayed organ donation decisions were not monitored because <50% of participants provided information required to check registration status or track removals, and NY State forbids checking the registry unless patients are candidates for imminent donation.

Another strength is that the videos presented compelling stories with real testimonials (Rodrigue, Fleishman, Vishnevsky, Fitzpatrick, & Boger, 2014; Singhal et al., 2004). Nonetheless, other production choices may have limited impact. One study suggested including a pediatric transplant recipient is superior to showing an adult who is waiting and that an uplifting ending is superior, as shown in our generic video (Rodrigue et al., 2014), while another study showed both uplifting and sad content are efficacious (J. T. Siegel et al., 2008). We designed the tailoring algorithm solely based on organ donation beliefs. Tailoring based on other attributes (e.g., emotional valence, types of spokespersons) might be more relevant (Rodrigue et al., 2014). Only 38% of eligible participants enrolled in the study when approached. Although attrition might have biased estimates towards increased registration, registration rates were less than expected.

## Conclusions

Neither targeted nor tailored videos differentially affected immediate registration among Black men in BOBs; however, targeted and tailored videos improved ODWS and were perceived more favorably than the generic video. Impact of tailored videos was no different from the targeted video, but when any content was removed during tailoring, outcomes trended towards being more favorable. Future work will report qualitative reasons why participants made their decisions from free text responses and data collected from semi-structured interviews conducted with a subset of these participants. Knowledge gained will inform improvements in video production and whether videos might be distributed via large-screen TVs in barbershops or social media with mobile applications.

## Data Availability

All data produced in the present study are available upon reasonable request to the authors, after this manuscript is accepted for publication.

## Abbreviations

CI: confidence interval;
DMV: Department of Motor Vehicles
BOBs: Black-owned barbershops
ODBI: organ donation belief index
ODWS: organ donation willingness stage of change;
OR: odds ratio

