## Supplements for "Targeting Versus Tailoring Educational Videos for Encouraging Deceased Organ Donor Registration in Black-Owned Barbershops"

### SUPPLEMENTAL INFORMATION

#### A. Educational-Entertainment Expertise and Expanded Methodology

The educational-entertainment expert is an Emeritus Associate Professor of Education, Communication, and Technology whose career spanned >40 years. The expert's research focused on the design and development of video-based learning environments informed by progressive, cognitive, and constructivist views of learning and instruction. The principal investigator attained a Master of Education, Communication, and Technology with the educational-entertainment expert chairing the thesis committee. Together, they produced and evaluated educational media about organ donation for >10 years. The producer is a Clio and Shorty Award winner, former Executive Producer of MTV World, and a former United Nations Global Accelerator Delegate, who has a track record of teaming with leaders in the medical community to fuse healthcare messaging with quality production and creativity.

Educational-entertainment models grounded in cognitive science, education, and social psychology, informed the production of videos distributed to increase deceased organ donor registration (A. Singhal, Cody, Rogers, & Sabido, 2004). Social cognitive learning theory suggests "modeling" realistic situations, social relationships, and attitudes with character portrayals of both careless and healthy behaviors and their consequences (Bandura, 1974, 1986, 2001; Blandin & Proteau, 2000; Olson & Bruner, 1974; A. Singhal et al., 2004; Slater, 2002; Vygotsky, 1978). Cognitive dissonance suggests media designs that juxtapose multiple opposing perspectives induce cognitive conflict, thereby heightening engagement and problem-solving (Elliot & Devine, 1994; Festinger, 1957; Harmon-Jones & Mills, 1999). "Anchored instruction" proposes portraying meaningful problematic social situations presented with multiple perspectives and alternative solutions through character interactions with unresolved endings that challenge viewers to explore problems further (Bransford, Sherwood, Hasselbring, Kinzer, & Williams, 1990; CTGV, 1990, 1993). These models are consistent with principles of the "docu-drama" genre and audience-centered media designs (A. Singhal et al., 2004; Sood, Menard, & Witte, 2004). In these approaches, confrontation generates emotions as motivating forces driving characters' actions; this functions to induce in viewers empathic responses and the conceptualization of problems and their resolutions as if occurring in their own lives (Kincaid, 2002). Like the models grounded in cognitive science, education, and social psychology, these models encourage viewers to live vicariously through entertainment media, recall prior experiences and use their imagination for conceptualizing story messages which have promise to modify behavior (Sood et al., 2004).

Guided by these principles, the production team casted experts and protagonists and drafted story boards, with content prioritized to address beliefs and misconceptions identified from the formative interviews. Video screening was informed by the IIFF model that postulates successful organ donation interventions provide 1) Immediate complete registration opportunities, 2) Information, 3) Focused engagement, and 4) Favorable activation (Alvaro, Siegel, & Jones, 2011; Siegel et al., 2016).

**B. Formative Research**

**Methods.** We conducted an exploratory qualitative study among Black clients in four Black-owned barbershops throughout Harlem, NYC from January-June 2014. Participants included BOB clients, who self-identified as Black or African American men, age 18 years or older, who live in NYC at least 28-days per year – to ensure participants were expressing views shared among the NYC Black community, and were fluent in English. Owners received a recruitment letter detailing the study purposes and their commitment. None refused. Owners determined the shop space for video interviewing that would not interfere with business operations while ensuring participant privacy. These designated spaces allowed the research team to conduct the interviews privately, without influence from shop owners and/or patrons.

The affiliated school of medicine's institutional review board approved the formative study. None of the study team members asked participants to register as an organ donor. To minimize partiality, our investigators comprised a multidisciplinary ethnically diverse team including a Black male research coordinator, who supervised all interviewing, but did not patronize any of the BOBs and knew none of the participants. A PhD prepared registered nurse with extensive expertise in qualitative methods trained the two research coordinators in data collection, interviewing, and coding. A Black male primary care physician from the New York Metropolitan area, who previously conducted qualitative research and health promotion studies with Black men in BOBs, facilitated the community partnerships and verified the coding scheme.

The framework for modeling individual decisions to donate organs posthumously proposed by Radecki/ Jaccard (1997) provided a structure for developing interview questions and qualitative coding (Radecki & Jaccard, 1997) (Tables S1 & S2). Based on reasoned action, the model describes individual and family factors that influence organ donation decisions and postulates that beliefs comprise an attitude that, in turn, leads to (or detracts from) willingness to donate.

To maintain credibility of the data, the same interview questions were posed to all participants. Questions ascertained participants' beliefs about organ donation and preferred ways for conveying information in educational videos (Table-S1). Screening demographic questions included age, sex, self-identified race and ethnicity, education, and religion. Participants then filled out the consent form and subsequently were interviewed. All interviews were audio and video recorded. Audio recordings were transcribed verbatim, with identities removed. To capture important nuances in communication, field notes were analyzed and video recordings reviewed for nonverbal behaviors.

Three team members used ATLAS.ti Version 7 (Berlin, Germany) for coding and analysis. Two trained research coordinators used frequency a-priori and in-vivo codes. The qualitative expert coded the same transcripts, checking for omissions, disagreements or inconsistencies. Analytic memos were discussed and as coders worked together to merge, delete, or rename codes when appropriate. Inter-coder agreement was monitored to realize a 80-90% minimal benchmark (Saldaña, 2013). During the interview, coding, and interpretation phases, research team members employed the peer review process to minimize bias. An audit trail was documented and reviewed to assist anyone wanting to replicate the study.

Barbers introduced potential participants to the research team, or they were approached directly, after information was provided information about the study followed by requests to enroll. Participants' organ donation registration choices were determined by administering the previously validated Organ Donation Willingness Scale [ODWS] (Boey, 2002; Parisi & Katz, 1986). The question, "How willing are you to sign up for organ donation?" allowed for six choices: "I do not want to sign up"; "I don't think I want to sign up; "I may sign up"; "I would like to sign up"; "I definitely would like to sign up"; I already signed up." Participants were grouped into those who were not interested (choices 1-2), those who were undecided (choice 3), and those who registered or planned to register (choices 4-6). Purposeful sampling was used to recruit equal numbers of Black men in each group. Once analytical saturation was reached, no one was added to that group. Every man who completed an interview received a free haircut valued at $25.

**Results.** Interviews were conducted with 79 men in four BOBs and averaged 22 minutes (range 10-35). After discarding two interviews due to equipment malfunction, 77 were retained: 27 registered or planned to register, 25 were undecided, and 25 were not interested in donating their organs posthumously. Participants who registered or planned to register were older, had more than high school education, and were not religiously observant (Table S3). Codes were then grouped under the Radecki-Jaccard framework (Table S2).

***Religion: "Your spirit will live on even without one of the organs"***

Participant undecided about organ donation.

None of the men justified their beliefs about organ donation based on specific religious dictum whether Muslim, Christian or Jehovah Witness. More than half the men said they regularly attended services. Religion provides a moral framework for their daily lives, and they cited their priests, ministers, imams or elders as respected leaders; however, their decisions were justified by other factors. "It's up to you to donate and not the church," said a man who had already registered. Men often discussed the greater importance of their 'spirit' after they died. As a Catholic man who registered noted, "I believe my spirit will leave my body and my body remains here. Once I enter the Kingdom of Heaven, I don't need my organs in physical form because I'll be spiritually with God." One Muslim man who would not register spoke about having his organs intact when he is buried.

***Culture: "Our people, my people, the Black people…a lot of have-nots, and they don't have the resources to get these donations"*** A man undecided about organ donor registration.

Cultural opinions about race dominated discussions. "Well, why are we not stepping up to the plate more? Other races are stepping up to the plate and we need this more. So, let's do this for ourselves. Let's help each other," replied a man who has already registered. Others cited race as a reason not to register. As a registered donor said, "There's a checkered history with the medical establishment and Black Americans. For good reason, I think there is skepticism."

> ***Knowledge: "I guess I would have to understand what the process would be. Would they take it out directly after I pass away? Will I still be able to be in a casket and everything?*** A man undecided about donation.

Even Black men who registered or plan to register as organ donors felt uninformed about donation practice. They often expressed, "I don't know exactly," or "I think I'd have to sign something," when asked about the process. Men who already registered mentioned donor cards and registration through the Department of Motor Vehicles (DMW), but no one was sure where registration lists were stored.

Respondents who are undecided or not interested expressed fear about the registration process. They commonly voiced three fears: being robbed of organs before they died, receiving less-aggressive medical care if physicians or emergency medical technicians knew they were organ donors, and not being eligible to donate because of habits and choices. "I know a friend that donated - he was in a bad car accident. They really didn't try to save him because he was a donor," stated a man uninterested in registering. Others revealed concerns about body disfigurement. A young man was concerned about his looks at his funeral, "Am I going to look sunk, are they just going to embalm me with so much fluid?"

Money was the most common reason motivating their registration decisions. "I do believe …that those who are more fortunate do not have to wait as long as those who are less fortunate because cash rules the world," said a man who registered despite his misgivings. When race and money codes were juxtaposed to assess their relative influence on donation process, money outweighed race, "To be honest, I think if I'm a Black guy and there's another Black guy beside me and I have more money than that person…even if I'm the bad person and he's a good person, I'll be most likely to be the next person in line. It doesn't really matter. It could be like a rich Black person compared to a poor white person. They're going to take that guy over just because of money wise," said a man not interested in registering. Another man who would not register believed professionals are not immune from wanting undeserved financial compensation. He said, "If that man is able to pay what that doctor wants, most doctors would make it [the donation] happen. Most hospitals will make it happen." Of note, we often dual-coded quotations related to the perceived impact of race and financial status on organ donation within both culture and knowledge domains given their inherent congruence, leading to merging these domains for video content purposes.

***Altruism: "Yes I'll donate because somebody's given me life. So, I want to give somebody else life."*** A man who registered.

Quotations reflecting altruism were closely related to culture and normative beliefs. Men across the willingness spectrum desired to save lives. Registered men or those willing to become donors were firmest in their altruism. "It's one of those things where if you can help someone out, why not? It's not like you're dying, you're dead. Why not help a life? It just makes sense to me," said a donation supporter. Men also spoke about being the recipient of a stranger's kindness and enjoying the feeling of knowing you are helping others. Even those men who said they would not register spoke about helping others as a noble act. As a one man said, "I would make it to a point where I don't really care about the race of that person...I'm gone from this world…as long as somebody gets it [my organ] to stay alive." His decision not to register was influenced more by his god-brother who died while waiting for a kidney.

Participants also expressed conditional altruism to ensure that recipients were worthy of their gift and that their organs went to someone in their community. "A man said he would not register until, "…there is a Black organization that just donates to Black people in general." Some men indicated a family member or a child in need would motivate them to donate while others stated they would donate only with assurances that their donation went to someone with no history of substance or alcohol misuse. Others expressed consternation if someone they considered undeserving possibly received their organs. "Do I have a choice to say who? I don't want a pedophile to have my heart. I feel like if it's my heart, I should have the right while I'm still living or my partner should have the right to decide who should have my organs," said a man not interested in registering.

***Normative: "Generally speaking, I think my family is in favor of organ donation. We're pretty progressive, you know?"*** A registered man.

Men in favor of donation or thinking about registering stated that they believed family members supported donation; five spoke about family members or friends who needed an organ transplant. A man planning to register talked about a family member who is on the waitlist for kidneys. He stated, "My family is kind of excited about the fact that s/he may have this opportunity." One man spoke about growing up in the South; he felt "It was southern hospitality, everybody did it [donates]." A man wanted to donate a kidney to his ailing family member, but sadly, his screening profile was not compatible. After this experience, he wished to register as an organ donor posthumously for someone else in need. Men's thoughts about family crossed into beliefs about altruism because more than half the participants said they wanted to donate organs to a family member. "If anything were to happen to my children, my grandchildren, and if something happens to me and they need it, they can get it," stated a man who may register.

> ***Encouraging organ donation with video education: "People are a little skittish about death, so we don't want to hear everything, every detail…I want to donate, and I just want to know what the process is."*** A man who may register.

All participants were queried about whether an educational video would encourage Black men to donate; and if so, what content would be most encouraging. Most participants believed viewers would be motivated to register if the video content included information about the organ donation process along with information about the low rate of registration among Black men. Some believed messaging should be positive, short and somehow emotionally touching. A man who registered stated, "I would point out the experiences and the lives that changed because of organ donation. Some people have lived because they got an organ and (some) were mothers and fathers…Everyone has a mother, and they love their mother and father dearly, so that could be something that would make them give more of themselves." Others believed showing the plight of those in need would be more effective. "If I see the type of pain that that person is going through late night when that pain really strikes you like a toothache and if I see those things, if I see the pain those people are going through, the hurt that they have, that'll touch them," said an uninterested man.

Participants suggested that well-known celebrities or well-regarded spokespersons and community leaders would make strengthen messaging. "I think people relate to celebrities," said one man. Some unwilling participants even suggested they would donate their organs to celebrities, as a man, "If Beyoncé told me to give her [an] organ, I would give her my organ right now!" While one participant believed politicians, secular and religious leaders are the "the voice in people of color's community, others disagreed about including a celebrity, as a uninterested man said, "I don't think it's a celebrity that can convince people…If you see a parent or somebody who had to let their son die…then I think people would feel that more than seeing a celebrity."

**Translating findings into video production.** Video content was informed from the voices of these 77 clients recruited from four BOBs. Those willing to donate their organs posthumously shared beliefs that: 1) their religion supports organ donation; 2) their physical body is not required for their afterlife; 3) the organ donation system prioritizes those in greatest need; 4) organ donation is the greatest gift one can give to another; and 5) family and friends would support their decision and share their beliefs (Arriola, Perryman, Doldren, Warren, & Robinson, 2007; DuBay et al., 2014; S. E. Morgan, Harrison, Afifi, Long, & Stephenson, 2008; S. E. Morgan & Miller, 2002; S. E. Morgan, Stephenson, Harrison, Afifi, & Long, 2008; Newton, 2011; Purnell et al., 2011; Siminoff, Burant, & Ibrahim, 2006). Those unwilling to donate believed that: 1) their bodies need to be intact for the afterlife; 2) doctors prioritize organ donation over saving lives of registered organ donors; 3) organ transplants are reserved for the wealthy; and 4) their family and friends are opposed to organ donation. Among our participants, the opposition to donation was further strengthened by beliefs that non-Latino whites have priority for transplants with Blacks donating organs without reciprocity (Arriola et al., 2007; Callender, Bayton, Yeager, & Clark, 1982; DuBay et al., 2014; Hinck et al., 2016; M. Morgan, Kenten, Deedat, & Donate Programme, 2013; M. Morgan, Mayblin, & Jones, 2008; S. E. Morgan, T. R. Harrison, et al., 2008; S. E. Morgan, M. T. Stephenson, et al., 2008; Newton, 2011; Purnell et al., 2011; Siminoff et al., 2006). Those unwilling or undecided might be swayed to register for organ donation if they could direct their organs to those from their same race/ethnicity and community or to those deserving of their organs, so that persons who damaged their organs carelessly or have poor character are excluded from their act of altruism (Arriola et al., 2007; DuBay et al., 2014; S. E. Morgan, M. T. Stephenson, et al., 2008; Newton, 2011). Also, interviews revealed secularization, so that religion seemed to play a lesser role in organ donation decisions and that money supersedes race when the medical system determines who receives transplants.

These results informed content organized by a modified version of the model for individual decision to donate organs posthumously proposed by Radecki/ Jaccard (Radecki & Jaccard, 1997). Given the strong correlation of codes for culture and knowledge, these constructs were compressed into a shared culture/knowledge domain. Therefore, video content included religious beliefs (for those with faith-based decision making), with more focus on addressing mistrust with the donation system – specifically how financial status impacts donation - and physician practices surrounding resuscitation of organ donors, altruism, and normative content.

While the content surrounding well regarded and celebrity spokespersons was informative, unfortunately budget constraints precluded hiring such persons for the video. This decision was also justified on theoretical grounds, as entertainment education theory states that celebrity spokespersons, while impactful, are subject to individual opinions about the spokespersons and also unpredictable events in that spokesperson's life that may negatively change public perceptions about their character and trustworthiness; furthermore, spokesperson popularity is time dependent, so inclusion impacts the longevity of the videos (Arvind Singhal, 2004). Some participants stated that celebrities would not be meaningful spokespersons, further justifying this decision.

Informed by these findings, the documentary style video focused on a Black man, James, as he struggles to support his family while he waits for a kidney transplant; the video also included uplifting scenes with his partner and stepdaughters. To balance participant preferences of ending choices, the story ends unresolved when James's states, "Hopefully someone somewhere will help me. I just patiently wait and wait." Hence, viewers are left to consider the outcome they hope for, expect, or fear for James and the consequences. This approach, presenting an unresolved story, aligns with literature on how best to encourage learning from video and, in particular, education video stories (Arvind Singhal, 2004). A complete description of the video, *James's Story,* is in the main documents.

**C. Data and Safety Monitoring Board and Community Advisory Board Descriptions/Roles**

The Data and Safety Monitoring Board comprised a statistician with extensive experience in data and safety monitoring with specialization in community-based research conducted among vulnerable populations, a renal transplant nephrologist, and a bio-ethicist. The board was available to discuss any adverse events (e.g., persons who were found to be psychologically unfit to participate either from initial screening or during participation) and to review study progress annually to determine whether the study should end early. Originally there was a plan to conduct interim analyses, but upon review, consensus was that important information would be learned from analyzing secondary outcomes, even if the primary outcome, increased registration, was not realized.

The community advisory board was founded and chaired by a Reverend who served as Rector of St. Philip's Episcopal Church in Harlem. After sadly passing away in March 2015, a Pastor from the North Presbyterian Church and Associate Professor of African and African American Studies specializing in "criminal injustice" agreed to serve as its new chair. The rest of the committee included four barbershop owners and two community representatives/activists, one of whom was female. The age range of members was 40-55. Members either lived in New York City or had businesses located within Harlem and/or Brooklyn. The project manager selected members because of their deep ties to Black communities where recruitment was conducted. Recruitment followed a snowball sampling approach, with the project manager, who previously worked with these same barbershops on preventive health endeavors, leading recruitment for barbershop owners and activists, while the Interfaith Center of New York assisted with recruiting the chairs.

The community advisory board met annually and as needed informally, to advise on study design, recruiting, and video production. Some board members were owners of shops where recruitment occurred, and hence were actively engaged in the study. Prior to distributing the videos, the project was formally presented to the board and feedback was solicited from members. Overall, the members were enthusiastic about the videos and the potential for encouraging registration. The main feedback was to end the video with James, the protagonist, lamenting about his waiting, so that scene would have a lasting impression for viewers to contemplate. The targeted and tailored videos were modified accordingly.

#### Supplemental Tables

##### Table S1: Semi-structured Interview Guide

| # | Questions |
|---|---|
| 1 | What is your opinion about organ donation knowing that it can save lives? |
| 2 | We have talked to different religious institutions about organ donation. You are described as X religion. We know that X religion is in favor of organ donation. What do you think about it? |
| 3 | If you, a member of your family or a friend would need an organ like a kidney for example; What do you think are the steps to follow for a doctor to get an organ? |
| 4 | If you want to donate your organs after you die, what do you think you would have to do to make the donation happen? |
| 5 | How do you think doctors decide who gets an organ transplant and who doesn't? |
| 6 | Why didn't (did) you sign up for organ donation? What would make you change your mind? |
| 7 | What do you think your relatives and / or friends think about organ donation? |
| 8 | If we showed a brief video about organ donation, do you think it would help people decide to be organ donors? Why? Why not? |
| 9 | What would you include in a video to get people to register to be an organ donor? |
| 10 | Who (e.g., community leaders, religious, etc.) should be included in this video to speak on organ donation? |
| 11 | Is there anything we did not ask that you would like to add? |

##### Table S2: Formative Study Demographics Stratified by Organ Donation Willingness

| | Willing (27) | Undecided (25) | Unwilling (25) | P value |
|---|---|---|---|---|
| **Age** | 45 (32, 51) | 31 (25, 51) | 31 (23, 50) | |
| **Ethnicity** | | | | 0.564 |
| Non-Latino | 25 (93%) | 21 (84%) | 21 (84%) | |
| Latino | 2 (7%) | 4 (16%) | 4 (16%) | |
| **Highest Level Education** | | | | 0.069 |
| High School or Less | 5 (18%) | 10 (40%) | 12 (48%) | |
| Greater than High School | 22 (82%) | 15 (60%) | 13 (52%) | |
| **Religion** | | | | 0.011 |
| Christian | 16 (60%) | 20 (80%) | 11 (44%) | |
| Other | 6 (22%) | 4 (16%) | 13 (52%) | |
| Not | 5 (18%) | 1 ( 4%) | 1 ( 4%) | |

All participants self-identified as men from Black, African American, or Afro-Caribbean/West Indies heritage. Median / IQR reported for continuous data; raw number and % reported for categorical data. For religion, Christian category includes self-identified Catholics, Protestants, and Christians. Other self-identified religious categories included Islam, Jewish, and Other. Not Religious included those who refused to answer or were agnostic.

**Table S3: Representative Quotes by Willingness to Register for Organ Donation**

| BEHAVIORAL CONSTRUCT | REGISTERED/WILLING N=27 | UNDECIDED N=25 | NOT INTERESTED N=25 |
|---|---|---|---|
| RELIGIOUS | After death, my body isn't really like that sacred. It's just..my soul and spirit, so having something just rot in my body for no reason when I could actually just contribute to someone's life immensely, I guess that just makes sense being Catholic that I would do that… | My feeling used to be that if God made me this way and I'm supposed to go to heaven or whatever my eternity is supposed to be, right, I should have everything with me. What should I do? | All our organs should go back with us to the grave. |
| CULTURE[@] | Because as black men…we tend to not live a long life. Rather if we go, we could save another life…If you're dead and gone, why not help a life? | Organ donation is not really something that's like [discussed] - especially in the black community. Donating your bread maybe, but like your organs, nobody would pressure you to do that or anything like that. | So if someone has a problem because that person is light skinned or this person is dark skinned and they're not going to give that person my organ because he's this…you can't blame them because that's the type of world we live in. |
| KNOWLEDGE[@] | I do believe in certain cases those who are more fortunate do not have to wait as long as those who are less fortunate because cash rules the world. I do believe that because a heart isn't black or white. A lung isn't black or white. I don't think it [money] should make a difference. | I'm kind of on the fence. I guess I would have to understand what the process would be as far as would they take it out directly after I pass away? Then when I get buried, will I still be able to be in a casket and everything? | I used to be an organ donor but I took myself off. Reason is being, I didn't know enough information about organ donation… |
| ALTRUISTIC | The reason I became an organ donor is because I had a good friend that I worked with for ten years and her [partner] needed a kidney. And, it was through that information that I realized, you know, if I pass away or if my kidney matched somebody... it should be known that that's ok. So, that's, you know, I've done it all my life. | I like to help people. Don't do it because you're expecting something back. You do it because that's the right thing to do. That's how I like to live. | I feel like maybe I'll be able to save a family member better than a stranger…I'd keep it tight for that. Hopefully my organs last long enough for me to be well until my old age but if I could help a family member, I'd do that before a stranger anytime. |
| NORMATIVE | That stems from like my [family member] being a nurse [for 25 years]…My [family member] needed a kidney so I guess I've witnessed it firsthand…luckily for her that she found a match, but I know that other folks aren't as fortunate. | My [family member] was African and nobody was talking his organs. His organs was his organs. | I am not going to give an organ of mine unless I discuss it with my family a little bit more. It will give me feedback on what I do. |

N – sample size, [@]Culture and Knowledge domains were highly correlated and thus merged for the purposes of tailoring video content and measuring baseline organ donation beliefs.

**Table S4: Organ Donation Belief Index Questions – English and Spanish**

| How much do you believe each statement? |
|---|
| **Religion** |
| 1. It would be OK to donate my organs when I die |
| 2. My religion is for organ donation |
| 3. My religion tells people to donate their organs |
| 4. My religion allows me to become an organ donor |
| 5. I do not need my organs after I die |
| **Culture/Knowledge** |
| 6. People receive organs regardless of their race |
| 7. Rich people get organs the same way that I would |
| 8. Famous people get organs the same way that I would |
| 9. A rich person will wait as long as a poor person for an organ |
| 10. If I am signed up to be an organ donor a doctor will work hard to save my life |
| 11. Organ donation does not discriminate between races |
| 12. The system for deciding who gets the next available organ is fair |
| **Altruistic** |
| 13. Organ donation is the greatest gift I could give someone |
| 14. Organ donation would allow something positive to come out of my death |
| 15. Becoming an organ donor is the right thing to do |
| 16. Organ donation would allow people to remember me as a good person |
| **Normative - Perceptions of Family and Friends** |
| 17. My family is for organ donation |
| 18. My partner is for organ donation |
| 19. My friends are for organ donation |
| 20. I should talk about organ donation with my family |

Responses for items are provided below.

| |
|---|
| 1. Disbelieve a lot |
| 2. Disbelieve moderately |
| 3. Disbelieve a little |
| 4. Neutral Selection |
| 5. Believe a little |
| 6. Believe moderately |
| 7. Believe a lot |

#### Table S5: Organ Donation Willingness Scale

| How willing are you to sign up for organ donation? |
|---|
| 1. I don't want to sign up |
| 2. I don't think I want to sign up |
| 3. I may sign up |
| 4. I would like to sign up |
| 5. I definitely want to sign up |

#### Table S6: Intrinsic Motivation Inventory Questions – English and Spanish

| How much do you believe each statement? |
|---|
| **Interest/Enjoyment** |
| 1. I enjoyed watching this video about organ donation |
| 2. This video about organ donation was interesting. |
| 3. This video about organ donation was entertaining. |
| 4. This video about organ donation was engaging. |
| 5. This video about organ donation held my attention. |
| 6. I liked this video about organ donation. |
| 7. I would watch this video about organ donation again. |
| Value/Usefulness |
| 8. This video about organ donation may help improve health for Latinos. |
| 9. This video about organ donation could help improve health for Latinos. |
| 10. This video about organ donation will help improve health for Latinos. |
| 11. This video about organ donation is necessary for improving health for Latinos. |
| 12. This video about organ donation is useful for improving health for Latinos. |
| 13. This video about organ donation is important for Latinos to watch. |
| 14. This video about organ donation should be shown to other Latinos. |

Questions 7 & 14 were removed due to underperformance during calibrations.
Responses for items are provided below.

| |
|---|
| 1. Not at all/neutral |
| 2. Believe a little |
| 3. Believe moderately |
| 4. Believe a lot |

Not at all/neutral encompassed "disbelieve a little," "disbelieve middle," "disbelieve a lot," and "neutral" categories due to low frequency of unfavorable ratings.

**Table S7. Unadjusted Differences in Organ Donation Belief Scores Pre-Post Intervention Received ITT[1]**

| Organ Donor Belief Scores[2] | Generic N = 447 | Targeted N = 450 | Tailored N = 456 |
|---|---|---|---|
| **Intention to Treat** | **u (95% CI)** | **u (95% CI)** | **u (95% CI)** |
| Religion | 0.15 (0.09, 0.21) | 0.31 (0.23, 0.39) | 0.28 (0.22, 0.35) |
| Culture/Knowledge | 0.32 (0.25, 0.39) | 0.19 (0.12, 0.26) | 0.18 (0.12, 0.23) |
| Altruism | 0.28 (0.21, 0.35) | 0.34 (0.28, 0.40) | 0.26 (0.20, 0.33) |
| Normative | 0.18 (0.12, 0.24) | 0.25 (0.18, 0.32) | 0.21 (0.14, 0.28) |
| Treatment Effects | | | |
| Religion | 0.15 (0.09, 0.21) | 0.29 (0.22, 0.36) | 0.32 (0.25, 0.40) |
| Culture/Knowledge | 0.32 (0.25, 0.39) | 0.18 (0.12, 0.24) | 0.19 (0.11, 0.28) |
| Altruism | 0.28 (0.21, 0.35) | 0.33 (0.28, 0.38) | 0.21 (0.10, 0.31) |
| Normative | 0.18 (0.12, 0.24) | 0.25 (0.20, 0.31) | 0.15 (0.07, 0.24) |

N – number of participants within a treatment group; n – number of participants within each cell; u – mean; Std – standard deviation
Significant differences were realized for religion and culture knowledge scores, with the tailored group performing best for religion after adjustment for those who watched a video that was tailored (treatment effects). The generic videos trended towards increasing culture/knowledge scores by ~.15 logits.

[1]Scores measured just before viewing the assigned videos were subtracted from scores measured just afterwards.

[2]Means reported using multiple imputation on 20 repeated data sets sampled with replacement and combined using Rubin's rules. Std was adjusted for clustering at the level of Barbershops.

**Table S8. Unadjusted Treatment Effects Main Outcomes**

| | Generic N = 447 | Targeted N = 668 | Tailored N = 238 |
|---|---|---|---|
| | **n (%) [95%CI]** | **n (%) [95%CI]** | **n (%) [95%CI]** |
| **Registered[1]** | 43 ( 9.6) [ 7.0, 13.0] | 68 (10.2) [8.0, 12.9] | 29 (12.2) [8.5, 17.1] |
| **Took Brochure[1]** | 208 (48.1) [42.7, 53.7] | 362 (55.8) [50.2, 61.2] | 142 (61.5) [53.6, 68.8] |

#### Supplemental Figures

##### Figure S1: Pre/Post Organ Donation Willingness Stage of Change by Intervention (Intention to Treat Analysis)

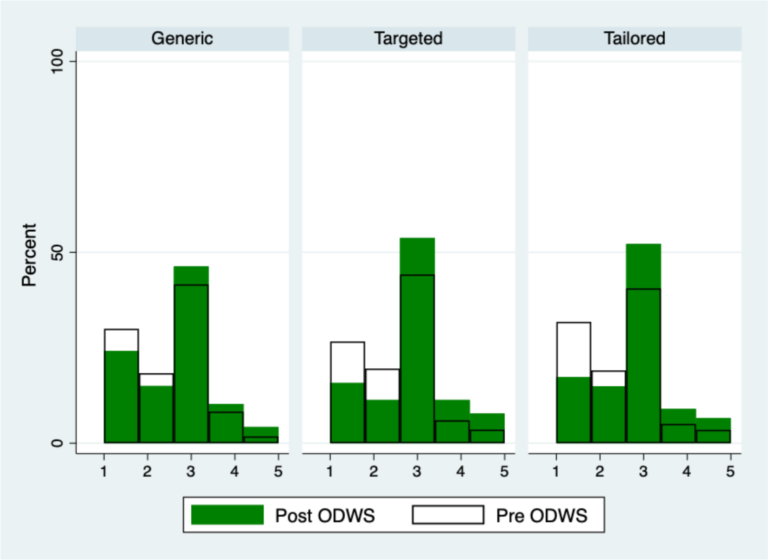


##### Figure S2: Pre/Post Organ Donation Willingness Stage of Change by Intervention (Treatment Effects Analysis)

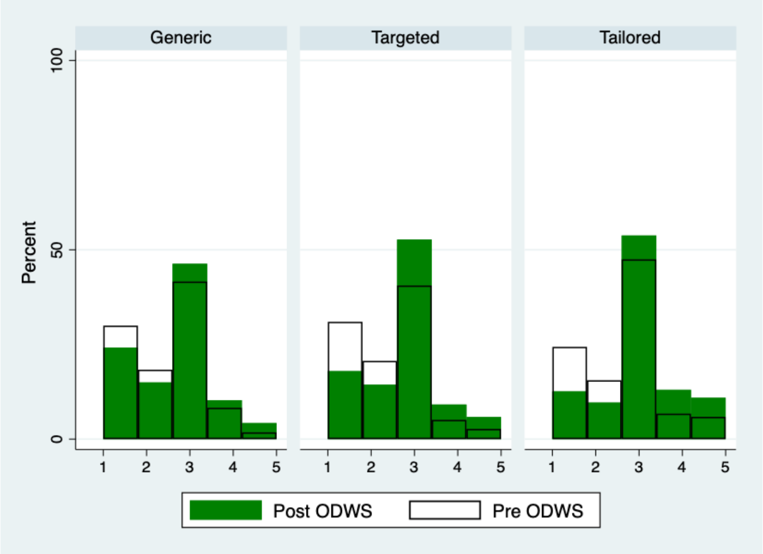


*Figures S1-S2 Summary:* Incremental increases in organ donation willingness stage of change are evident from pretest to posttest assessment. In the ITT analysis, the Targeted video seemingly had the greatest impact; however, when persons in the Tailored arm who saw the complete video, because of their response to the organ donation belief index (ODBI) scores within each construct, were moved into the Targeted video group, the remainder who actually watched a tailored video exhibited the greatest impact. Organ donation willingness stages include: 1) I don't want to sign up, 2) I don't think I want to sign up, 3) I may sign up, 4) I would like to sign up, 5) I definitely want to sign up.
